# Emergence of multiple SARS-CoV-2 mutations in an immunocompromised host

**DOI:** 10.1101/2021.01.10.20248871

**Authors:** Elham Khatamzas, Alexandra Rehn, Maximilian Muenchhoff, Johannes Hellmuth, Erik Gaitzsch, Tobias Weiglein, Enrico Georgi, Clemens Scherer, Stephanie Stecher, Oliver Weigert, Philipp Girl, Sabine Zange, Oliver T. Keppler, Joachim Stemmler, Michael von Bergwelt-Baildon, Roman Wölfel, Markus Antwerpen

## Abstract

Prolonged shedding of infectious SARS-CoV-2 has recently been reported in a number of immunosuppressed individuals with COVID-19. Here, we describe the detection of high levels of replication-competent SARS-CoV-2 in specimens taken from the respiratory tract of a B-cell depleted patient up to 154 days after initial COVID-19 diagnosis concomitant with the development of high mutation rate. In this patient, a total of 11 nonsynonymous mutations were detected in addition to the Y144 deletion in the spike protein of SARS-CoV-2.

Virus evolution studies revealed a dramatic diversification in viral population coinciding with treatment with convalescent plasma and clinical respiratory deterioration. Our findings highlight the urgent need for continuous real-time surveillance of genetic changes of SARS-CoV-2 adaptation alongside immunological investigations in patients with severely compromised humoral responses who may shed infectious virus over prolonged periods of time.

## Introduction

Since the end of 2019, the severe respiratory disease Coronavirus disease 2019 (COVID-19) [1] that originated in Wuhan, China, has developed into a pandemic with devastating effects. According to the John Hopkins University there are to this date (Dec. 22, 2020) over 77.4 million COVID-19 cases and 1.7 million deaths worldwide [2].

In December 2020, a new emergent variant of SARS-CoV-2 was reported in the United Kingdom [3] with a postulated higher infectivity and thus transmission rate [4]. Unique national and international collaborative effort has allowed sequencing data of full-length SARS-CoV-2 genomes to be shared immediately allowing both predictions of structural changes of specific viral proteins [5–8] important for immune recognition and the use of data for molecular epidemiology studies [9]. One of the possible sources for increased genetic diversity seen within SARS-CoV-2 may be individuals with prolonged viral infection due to underlying immunosuppression. Recently a number of single case reports have shown within host genetic diversity arising in the absence of immunological pressure and upon treatment with COVID-19 specific treatment in immunocompromised hosts [10][11]. In the context of the novel B 1.1.7 lineage a combination of mutations and deletions within spike were described in such a patient [10][11].

We have recently reported our observations with a cohort of six B-cell depleted patients with COVID-19 (BLD-2020-010058, under review). Within this study, we observed notable findings in one of those patients, who exhibited a protracted, eventually fatal clinical course characterised by the presence of high concentrations of replication-competent SARS-CoV-2 in the respiratory tract for more than 134 days. Here, we present analysis of the viral evolution of samples from this patient over time with increased detectable rate of mutation in comparison to two other B-cell depleted patients as well as the general population of SARS-CoV-2 infected individuals, thus suggesting a unique interplay of host and viral factors.

## Results

A woman (patient A) in her 70ies was hospitalised with COVID-19 and severe ARDS. She had been feeling unwell with worsening upper respiratory symptoms for two weeks prior to presentation requiring high flow oxygen on admission. On transfer to our center endotracheal intubation and ventilation were initiated and she was managed in our intensive care unit until her death 5 months later. Her past medical history was significant for follicular lymphoma that had been treated with a total of three cycles of standard chemotherapy cyclophosphamide, doxorubicin, vincristine and prednisone in combination with the B-cell depleting anti-CD20 antibody obinituzumab up to a month prior to presentation.

COVID-19-directed treatment initially included steroids as well as administration of convalescent plasma (Figure 1). Therapy with remdesivir was deemed unsafe due to existing renal failure and known hypersensitivity to sulfobutylether-β-cyclodextrin.

**Figure 1.**
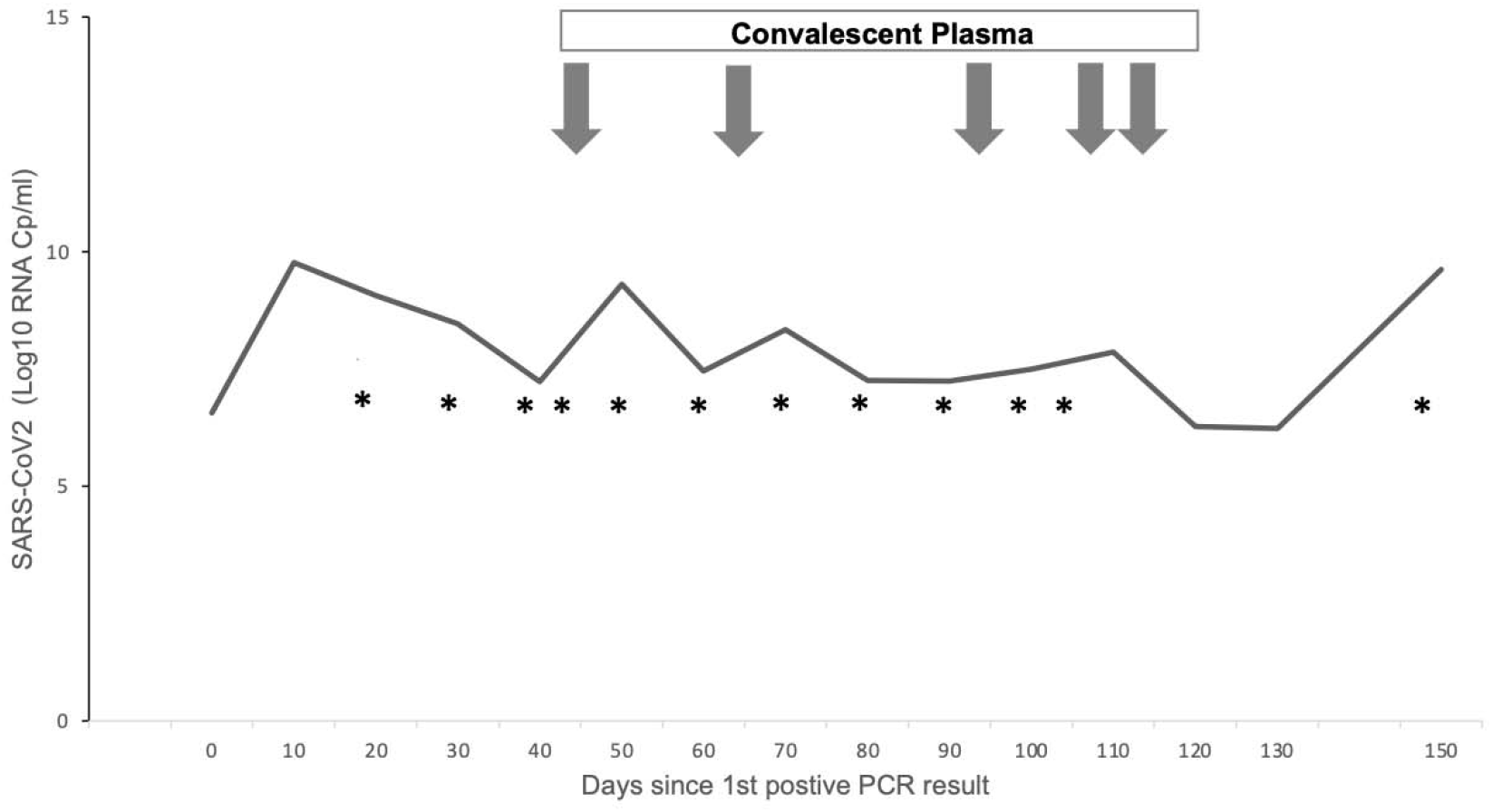
Time course of SARS-CoV-2 viral concentrations in endotracheal aspirates. X axis depicts time course from first positive PCR result, Y axis shows Log10 levels of SARS-CoV-2 RNA collected via endotracheal aspirates in Cp/ml. All specimens were positive for SARS-CoV-2 in culture. Arrows indicate administration of convalescent plasma. * indicates sequenced samples.

**Figure 2.**
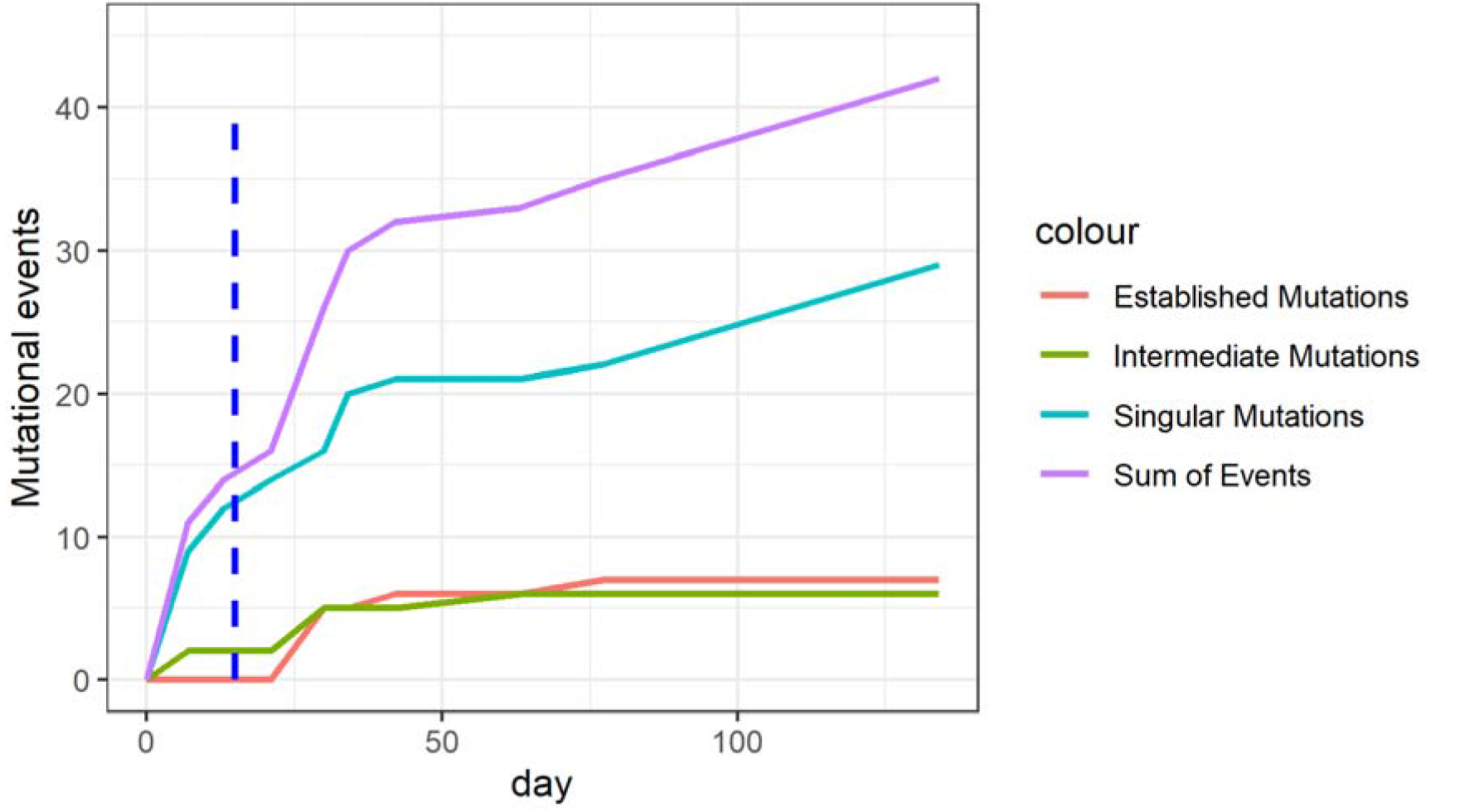
Mutational events over time grouped according to mutation classification seen for patient A. Dashed blue line shows the first administration of convalescence plasma.

Clinical upper and lower respiratory tract specimens obtained from patient A showed low Ct values and high viral concentrations throughout until date of death on day 156 following diagnosis (Figure 1). In keeping with this SARS-CoV-2 could be successfully isolated in Vero E6 cell cultures from all 16 patient samples examined in this study. Of note, the first samples submitted for viral cultures for patient A were on day 20, which although later in course of infection, we henceforth refer to as day 0.

At the same time we were also treating other B-cell depleted lymphoma patients within our unit (BLD-2020-010058, under review), two of these (patients B and C) protracted severe COVID-19 course with ARDS following nosocomial transmission of SARS-CoV-2 from a presumably common source. Eventually, both recovered fully and were able to be discharged from hospital care.

We were able to generate 16 high quality genomes of SARS-CoV-2 isolated from all three patients from successive time points (10 from patient A and 3 from patients B and C, respectively). All genomes were generated from samples with replication-competent SARS-CoV-2. The genomes hold at least 29685 nucleotides in length, which is more than 99% of the SARS-CoV-2 genome with a sequencing depth of usually more than 1000-fold (mean: 812-fold, min: 308-fold) (Table 1). Missing nucleotides are only located at the outermost ends.

**Table 1:** Overview of genomes metadata and summary of observed mutations in this study. For classification of mutations, please refer to Suppl_Data_File1.

| Day | Patient | Genome length | Genome coverage | ct-Value (RdRP) | Established mutations | intermediate mutations | Singular mutational events |
| --- | --- | --- | --- | --- | --- | --- | --- |
| 0 | A | 29685 | >1000-fold | 19.28 | 0 | 0 | 0 |
| 7 | A | 29978 | >1000-fold | 21.07 | 0 | 2 | 9 |
| 13 | A | 29873 | 617-fold | 16.77 | 0 | 0 | 3 |
| 21 | A | 29869 | 767-fold | 26.36 | 0 | 0 | 2 |
| 30 | A | 29853 | 400-fold | 26.60 | 5 | 3 | 2 |
| 34 | A | 29872 | 328-fold | 17.56 | 0 | 0 | 4 |
| 42 | A | 29890 | 308-fold | 25.15 | 1 | 0 | 1 |
| 63 | A | 29872 | 763-fold | 25.88 | 0 | 1 | 0 |
| 77 | A | 29901 | >1000-fold | 23.62 | 1 | 0 | 1 |
| 134 | A | 29898 | >1000-fold | 14.62 | 0 | 0 | 7 |
| 0 | B | 29901 | >1000-fold | 25,17 | 0 | 0 | 0 |
| 7 | B | 29873 | >1000-fold | 21,76 | 1 | 0 | 3 |
| 13 | B | 29865 | >1000-fold | 27,31 | 0 | 0 | 1 |
| 0 | C | 29900 | >1000-fold | 18,03 | 0 | 0 | 0 |
| 6 | C | 29901 | >1000-fold | 21,61 | 0 | 0 | 1 |
| 19 | C | 29888 | >1000-fold | 21,53 | 0 | 0 | 1 |

All mutations and mutational events are listed in Supplemental Data Table 1. Five mutations in total enable the grouping of all sequenced isolates into nextstrain-clade 20B (Old Nextstrain CladeA2a, Pangolin Lineage B.1.1, GISAID Clade GR). The observed mutations at nucleotide position 23403 (S-protein 614D>614G), 3037, 14408 and 241 belong to the most prominent European cluster. The deletion at position 28881 in the open reading frame of the N-protein was detected as well. In respiratory tract samples from patient A additional unique mutations were detected, for instance the mutation at position 19825 G>T (orf1b 6521V>6521L). This is up to now specific for sequences from the South of Germany sampled between 16 March – 11 May 2020, previously provided by the Max von Pettenkofer Institute in Munich, Germany (BY-MVP-0064/2020; 0073; 0118; 0219; 0283; V2012622; V2012612).

We also sequenced SARS-CoV-2 genomes isolated from patients B and C. Similar to patient A, they were diagnosed SARS-CoV-2 positive by PCR for 53 and 91 days, respectively. But both showed, in contrast to patient A, decreasing viral concentrations in respiratory tract specimens with replication-competent SARS-CoV-2 for only 14 and 20 days, respectively. COVID-19-directed therapy for all three patients consisted of multiple administrations of convalescent plasma. Sequencing of viral isolates from patients B and C demonstrated only 1-2 mutational events in 19 days, which is in accordance with the expected number of SARS-CoV-2 mutations. It should be stressed that this is a similar number of mutations observed in patient A within the initial 21 days. Patients B and C showed both the same mutational pattern supporting the hypothesis of a common source of infection. Detectable SARS-CoV-specific IgG could not be measured in either patients throughout, albeit in patients B and C transient low level antibodies were detected briefly following convalescent plasma treatment (Supplementary Data, Table 1).

In patient A sequences collected on day 30 (50 days following diagnosis) showed a notable increase in detectable mutations. Preceding this was treatment with two doses of convalescent plasma. Respiratory samples of patient A displayed an additional 5 mutations (one synonymous, two non-synonymous, deletion S protein H144 and one intergenic SNP), established at this time that could be tracked until day 134. Over time from day 30 onwards, another 11 additional detectable mutations accumulated (two synonymous and nine non-synonymous, Supplementary Data Table 2).

**Table 2:** List of non-synonymous mutations and deletions observed during this study of patient A. List comprises clone-specific mutations (n=8), as well as mutations seen during this study for this patient. For a full list of mutations, please refer to Suppl_Data_ Table1.

| gene | nucleotide | amino acid | remarks |
| --- | --- | --- | --- |
| **5'-UTR** | C241T | - |  |
| **Orf1a** | G558T | G98V |  |
|  | A2270G | K669E |  |
|  | G3564T | G1100V |  |
|  | A4230T | T1322I |  |
|  | C4241G | I1326V | Established mutation |
|  | C5178T | T1638I | Clone-specific mutation |
|  | A9741G | N3159S |  |
|  | C10116T | T3284I |  |
|  | A11451G | Q3729R |  |
|  | G12566T | V4101F | Clone-specific mutation |
|  | C12756T | T4164I | Established mutation |
|  | C14408T | P4715L | Clone-specific mutation |
|  | G19825T | V6521L | Clone-specific mutation |
| **S** | 21990-21993del | Y144del | Established inframe deletion |
|  | G22992A | S477N |  |
|  | G23012A | E484K | Established mutation |
|  | A23403G | D614G | Clone-specific mutation |
| **inter-orf** | G27201T | - | Established mutation |
| **Orf6** | GTT27214ATA | V5I |  |
|  | T27793C | F13S | Established mutation |
| **inter-orf** | TAAAATG 28270 TAATG<br>2bpdel | - | Clone-specific mutation |
| **N** | GGG28881AAC | RG203KR | Clone-specific mutation |
|  | A29247T | T325K | Established mutation |
|  | C29268T | T332I | Clone-specific mutation |

## Discussion

So far, only little detailed knowledge of immunosuppressed COVID-19 patients is available. Recent data has described significantly increased mutation rate and accumulation of these mutations within a short period of time in single case reports [10–12]. As SARS-CoV-2 was not only detectable but also cultivable from respiratory tract samples over such a long period of time, comprehensive sequencing data of SARS-CoV-2 could be generated from this single severely immunocompromised host. These sequences show an unusually large number of nucleotide changes and deletions in addition to an unusually high ratio of nonsynonymous mutational events. In our study, the ratio of non-synonymous to synonymous mutations is 3.25 and hence very high

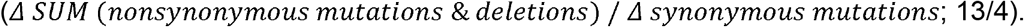

This is observed without any detectable SARS-CoV-2 specific antibody response by the host and after administration of convalescent plasma, similar to the findings described by Kemp et al [12]. The therapeutic efficacy of convalescent plasma is still under investigation in multiple clinical trials but so far has been disappointing [13]. Our data suggests that its use should be critically evaluated in patients with prolonged viral replication, where the effect of exerting external immunological selective pressure on a large viral population remains unknown. In particular, given our current incomplete understanding of the interaction between different hosts and SARS-CoV-2, convalescent plasma therapy could support the emergence of viral escape variants over time. This would further complicate treatment options and epidemic control.

With respect to the new emerging lineage B1.1.7 with its dominant deletion at position 21765-21770 (HVdel69-70) [4], our strain does not belong to this lineage. This specific deletion has already previously been observed and seems to be established independently at least two times (lineage 1.1.7 and 1.258), but in combination with mutation in the spike protein P681H (nt23604) and N501Y (nt23063), this seems to be unique at the moment [14]. In contrast to this, our samples show a deletion in the spike protein at Y144, which is not only observed in the emerging lineage B1.1.7 [4], but has also been described in a sample from 30 July 2020 in England (England/SHEF-C71F3/2020) [14]. As more detailed metadata of this strain is currently not available, one speculative explanation may be that these nucleotide changes were also generated in a chronically infected host.

Based on the assumption that circulating SARS-CoV-2 lineages accumulate nucleotide mutations at a rate of about 1-2 mutations per month [16], resulting in about 23,638 established mutations over year per genome [14], we would expect 8.64 mutations within 134 days, as seen for patient B and C (1.2 mutations in 19 days). However, for our index case, patient A, we could detect 16 mutations, which means more than a double (43.58 mutations per year) of the expected amount. Whether this occurs in the context of a quasispecies concept remains to be seen.

No additional SNPs affecting target sites of therapeutic agents like remdesevir were observed in our dataset. A number of monoclonal antibodies for treatment of SARS-CoV-2 infection are currently under evaluation and further work should elucidate the putative effect of these mutations on the target sites of these agents.

Our data have a number of limitations. They are based on an unusually long course of chronic infection with SARS-CoV-2 in a single individual, to our knowledge the longest published duration of infectious viral shedding since the start of the pandemic. Further we have no data on the neutralizing capacity of SARS-CoV-2-specific antibodies, including the applied convalescent plasma therapy, on the isolated viral populations. We assume little effect based on unaltered detectable viral concentrations measured from respiratory tract specimens throughout the course of infection. We also have no data on potential higher transmission capabilities conferred by the observed mutations. Enhanced appropriate infection control precautions during the prolonged care of this patient on our intensive care unit were sufficient and no nosocomial transmissions were observed.

In conclusion, we find that detailed characterization of the interaction between SARS-CoV-2 and the host in immunocompromised, particularly B-cell-depleted patients, provides a rare opportunity to follow viral adaptive evolution during the course of chronic infection. This will be crucial to understand the role of host-specific factors and help in the development of preventive and therapeutic agents. Further studies should therefore also specifically investigate the therapeutic efficacy of convalescent plasma therapy and protective effect of SARS-CoV-2 vaccines in immunocompromised individuals at increased risk of severe COVID-19 and fatal courses. We suggest that such studies should be done in collaboration between virology laboratories and clinical treatment centers to share findings in real-time and to translate them into therapeutic concepts as quickly as possible.

## Data Availability

All original data can be provided on request

## Ethical Statement

Patients are part of the COVID-19 Registries of the LMU University Hospital Munich (CORKUM, WHO trial id DRKS00021225). Clinical and routine laboratory data was prospectively collected within the COVID-19 registries and verified and complemented by individual chart review. Patient data were pseudonymized for analysis and the study was approved by the local ethics committees (No: 20-245). Written informed consent was obtained from each patient or authorized by proxy before any study related procedure.

## Acknowledgment

We greatly acknowledge all the colleagues working on the intensive care unit at the LMU Hospital in Großhadern Munich and the diagnostic team at the Bundeswehr Institute of Microbiology (IMB), for their dedicated and excellent work in nursing and diagnostics of our patients.

