## Supplementary Data for "Emergence of multiple SARS-CoV-2 mutations in an immunocompromised host"

**Supplemental Data**

|  | **SARS-CoV-2**  **IgG Titre** | **Day** |
| --- | --- | --- |
| **Donor 1** | 3.9 | 13 |
| **Donor 2** | 1.5 | 27 |
| **Donor 2** | 1.5 | 64 |
| **Donor 3** | 7 | 70 |
| **Donor 3** | 7 | 73 |

**Suppl Table 1.** SARS-CoV-2 IgG Titres of convalescent plasma treatments administered to patient A

### **Material and Methods**

### Measurement of SARS-CoV-2 antibodies

### For structural antibody testing, anti-SARS-CoV-2 IgG and IgA ELISAs were performed strictly according to the manufacturer's instructions (Euroimmun, Lübeck, Germany). Both IgG and IgA target a recombinant S1 domain protein (spike protein). The results are each expressed as a ratio calculated by dividing the optical densities (OD) of the sample by those of the internal calibrator provided. Samples were evaluated as suggested by the manufacturer as either nonreactive (ratio <0.8), borderline (0.8 ≤ ration ≤ 1.1), or reactive (ratio > 1.1). Both reactive and borderline results were classified as "reactive".

Viral culture and molecular identification

VeroE6 cells were inoculated at a dilution of 1:100 with an adsorption time of 1 hour at 37° C and shaken every 15 minutes. The cells were examined for cytopathic activity (CPE) every 24 hours. 72 hours after infection (h.p.i.), the supernatants were collected, clarified, aliquoted and stored at -80 ° C prior nucleic acid extraction. For  Illumina cDNA RNA-seq as well as screening by quantitative RT-PCR  (Corman et al., 2020), viral RNA was prepared from 200 µl of clarified virus stocks (3.000 rpm, 5 min) with the QIAamp Viral RNA Mini Kit (Qiagen, Hilden, Germany) and eluted in 50 µl RNase free H_2_0.

NGS Sequencing and phylogenetic classification

The extracted RNA was translated into cDNA using the SuperScript IV First-Strand Synthesis System (Invitrogen, Thermo Fisher Scientific, Dreieich, Germany). After performing second strand synthesis (non-directional RNA second strand synthesis module NEBNext Ultra II, New England Biolabs, Frankfurt am Main, Germany), a library was generated using the Twist Library Preparation Kit Twist Biosciences (San Francisco, CA, USA). In addition, a target enrichment step was added prior to sequencing on an Illumina MiSeq (Illumina Inc., Berlin, Germany). For this purpose, bait-set including SARS-CoV-2-specific capture baits ([Twist Respiratory Virus Research Panel](https://www.twistbioscience.com/getquote?Conversion_url=https%3A%2F%2Fwww.twistbioscience.com%2F), Twist BioSciences) were used according to the manufacturer's instructions and captured libraries were sequenced using Sequencing V2 reagent chemistry at 2x150 cycles on a Micro Flow Cell (Illumina Inc., Berlin, Germany).
Raw reads were in a mapping approach using bwa-aligner (Li, 2013)  V2.1 against the reference strain Wuhan-Hu-1 (MN908947.3). Variant calling as well as annotation were performed using FreeBayes Version v1.3.1-dirty (Garrison and Marth, 2012).

For this study we classified the observed mutations into four categories:

1. Clone-specific mutations observed for all of the sequences over the whole time of the study. Those mutations are of high relevance for use in phylogenetic studies.
2. Established mutations, which appeared during the study, were visible in at least two subsequent samples and stayed until the last sample.
3. Intermediate mutations, which appeared during the study, were visible in at least two subsequent samples, but disappeared until the last sample.
4. Singular mutational events, that are based on mutations, that were only present once in this study.

For phylogenetic classification, strains were submitted to a local copy of Nextstrain and clade assigning was performed using the implemented algorithms. Two genomes of patient A isolated from days 0 and 134 respectively were submitted to GisAID and were made publically available under AccNo EPI_ISL_732538 and EPI_ISL_732658. Submitted genomes of patient B and patient C are accessible under AccNo EPI_ISL_732533 and EPI_ISL_732560, respectively.
